# Investigating penetrance of severe combined immunodeficiency variants in an adult population cohort: implications for genomic newborn screening

**DOI:** 10.64898/2026.02.17.26346478

**Authors:** Imogen JH Grimwade, James Fasham, Caroline F. Wright, Leigh Jackson

**Affiliations:** Department of Clinical and Biomedical Sciences, College of Medicine and Health, University of Exeter Medical School, Exeter, UK; Department of Clinical Genetics, Royal Devon University Hospital, EX1 2ED Exeter, UK

**Keywords:** severe combined immunodeficiency, penetrance, genomic newborn screening

## Abstract

Severe combined immunodeficiency (SCID) is a heterogeneous, recessive disorder, associated with the onset of severe, recurrent infections in the first few months of life. SCID is fatal if left untreated, but outcomes can be significantly improved by prompt diagnosis and treatment, particularly prior to onset of infection. Consequently, SCID is already included in many newborn screening programmes around the world, as well as multiple international genomic newborn screening (gNBS) research programmes. However, there is a vital need to estimate penetrance of SCID variants in population cohorts, to mitigate the potential consequences of reporting low penetrance variants in a genotype-first gNBS setting. This study aimed to assess the penetrance and prevalence of these variants in the UK Biobank population cohort. Whole genome sequencing data from 490,640 individuals was used to interrogate 16 SCID genes for potentially causal variation. We identified 4206 carriers of single heterozygous pathogenic variants (∼1% of cohort), but only 6 individuals double heterozygous, homozygous or hemizygous for relevant pathogenic variants. 3 individuals would be expected to require further testing had they been identified by gNBS, suggesting that fewer than 1 in 100,000 newborns might require follow-up testing due to SCID variants. Following detailed variant curation, we were able to identify only 2 unaffected individuals likely to be harbouring biallelic pathogenic variants, potentially indicative of reduced penetrance. Nonetheless, SCID remains an excellent candidate for inclusion in gNBS studies, due its severity, clinical actionability and expected low false positive rate, although care should be taken when reporting hypomorphic variants.

## Introduction

Severe combined immunodeficiency (SCID) is a genetically heterogeneous immune disorder characterised by very low T-cell counts, which may occur alongside defects in other lymphocyte classes, depending on the genetic aetiology (1). This results in insufficient immune responses, typically leading to the onset of repeated, severe infections in the first few months of life, which are fatal if treatment is not commenced promptly (2–5). As a result, SCID is already included in numerous newborn screening programmes worldwide, using the targeted T-cell receptor excision circle (TREC) assay (6,7). Pathogenic variants in more than 15 genes are considered causal, with most cases inherited in an autosomal recessive manner, while a large proportion are X-linked recessive (1). The majority of typical SCID cases are caused by null variants, while atypical or ‘leaky’ SCID, is usually caused by hypomorphic variants in the same genes (1). SCID has an incidence of 1 in 22,819 - 127,000 live births, with this variability likely due to population differences, such as differing rates of consanguinity (8–10).

Due to its rarity and predominantly recessive nature, SCID cases often arise in the absence of known family history. Consequently, in geographies where SCID newborn screening is not yet routine, diagnosis is usually prompted by severe, recurrent infections in newborns, alongside low naïve T-cell counts (1). In the UK’s National Health Service, patients with suspected SCID will be offered single gene or whole genome sequencing (WGS), both to confirm the diagnosis and to inform immediate clinical management (1,11,12). The current standard of care for SCID patients is haematopoietic stem cell transplant (HSCT), which has the potential to be curative (2–5). However, neonatal diagnosis and prompt HSCT, especially prior to onset of infection, significantly improves survival, indicating that neonatal screening will be essential to improving outcomes for these patients (2–5).

As such, SCID and its various associated genes, are currently being assessed as candidates for genomic newborn screening (gNBS) in at least 27 research studies (13). gNBS programmes aim to use WGS to test apparently asymptomatic newborns for treatable genetic conditions, in particular those for which early treatment is advantageous (14). The ability of WGS to simultaneously screen for many conditions, provides an attractive prospect for expanding current newborn screening programmes, particularly for genetically heterogeneous conditions like SCID (14). However, there are vast challenges regarding the interpretation of genetic variants in an asymptomatic context, with little currently understood about the penetrance and expressivity of ostensibly pathogenic variants, in population cohorts (14,15). Since penetrance has historically been calculated from clinical cohorts, it is often overestimated, leading to uncertainty surrounding whether newborns who screen positive, will actually the develop the condition and if so, when (16,17).

The UK’s gNBS programme, the Generation Study, proposed including 16 SCID genes in its initial list of reportable conditions (13). Whilst some SCID genes are included in almost all gNBS programmes, others are discordantly represented (13). This raises the question of whether some individuals with SCID will be missed due to the exclusion of particular genes, or whether inappropriate inclusion of uncertain gene-disease pairs may lead to overdiagnosis. Ensuring that gNBS will not miss cases is especially relevant given that the measurement of TRECs in dried blood spot samples that are routinely collected, is highly sensitive and already used in many NBS programmes (6,7). gNBS research programmes will be crucial in providing insight into whether WGS will deliver an acceptable level of sensitivity and specificity for SCID screening, and how these parameters compare to TREC screening.

In addition to restricting analysis to established gene-disease pairs, gNBS specificity can be achieved by ensuring that all conditions selected for gNBS programmes are highly penetrant, an essential prerequisite for inclusion (14,15). The penetrance of pathogenic variants in the majority of the 16 SCID genes included in the Generation Study has been estimated at over 80% (18,19). However, given that penetrance calculations have been derived from small clinical cohorts (18,19), this is likely an overestimate (16,17). In addition, the estimation of penetrance of variants in less well characterised SCID genes, has been precluded by the small numbers of affected individuals. As such, there is uncertainty surrounding the penetrance of SCID variants in the population, and therefore how these will be interpreted in the context of asymptomatic genome screening.

There is a vital need to address these uncertainties surrounding penetrance in population cohorts, to enable accurate variant interpretation and risk prediction in gNBS programmes. In this study, we aimed to assess the penetrance and prevalence of SCID variants in the UK Biobank (UKB) population cohort, to provide more insight into how these variants should be interpreted if identified in asymptomatic newborns.

### Subjects and methods UKB cohort

The UKB is a population cohort of 501,936 individuals, recruited between 2006 and 2010, at which point they were aged 40 to 69 years (20). On recruitment, participants answered questionnaires concerning lifestyle and medical history, as well as provided physical measurements and biological samples (20). These samples have since been used for assays such as blood cell counts and proteomics, the latter providing data on the expression of proteins encoded by certain SCID genes. In addition, hospital episode statistics and GP clinical event records are available for 450,852 and 229,839 participants, respectively, enabling extensive phenotypic profiling (20).

In addition to exome sequencing data, WGS has now been performed on blood samples from 490,640 participants, making up 98% of the cohort (21). Short-read WGS was carried out using the Illumina NovaSeq 6000 sequencing platform, followed by alignment to the GRCh38 reference genome and variant calling using Illumina’s DRAGEN v3.7.8 pipeline (21).

### Classifying variants

WGS data was used to investigate *IL2RG*, *ADA*, *RAG1*, *CD3D*, *IL7R*, *JAK3*, *RAG2*, *NHEJ1*, *PTPRC*, *DCLRE1C*, *CD3E*, *CD3G*, *CD247*, *AK2*, *CORO1A* and *ZAP70* for potentially pathogenic variation. Interpretation and classification of variants was carried out consistently for all 16 genes, as detailed in Figure 1. Variants were annotated using Ensembl’s Variant Effect Predictor, with only the annotations against the MANE Select transcript for each gene, retained. Variants with ≥1-Star ClinVar classifications of Benign, Likely Benign or Uncertain Significance were excluded (22). Conflicting classifications were retained only if the majority were Pathogenic/Likely Pathogenic. With the exception of those annotated as splice acceptor or donor variants, intronic, synonymous and missense variants were excluded, unless they had previous ClinVar ≥1-Star Pathogenic or Likely Pathogenic classifications (22). Predicted loss-of-function (pLOF) variants were retained, including frameshift, nonsense, splice acceptor and splice donor variants, except those found in nonsense mediated decay escape regions. These were determined as the first 150 base pairs and the last exon, as well as the last 55 base pairs of the penultimate exon (23). In exception to this, pLOF variants in nonsense mediated decay escape regions were retained if variants had Pathogenic/Likely Pathogenic classifications or if Pathogenic/Likely Pathogenic variants had previously been identified in these regions, as is the case in *IL2RG*.

**Figure 1.**
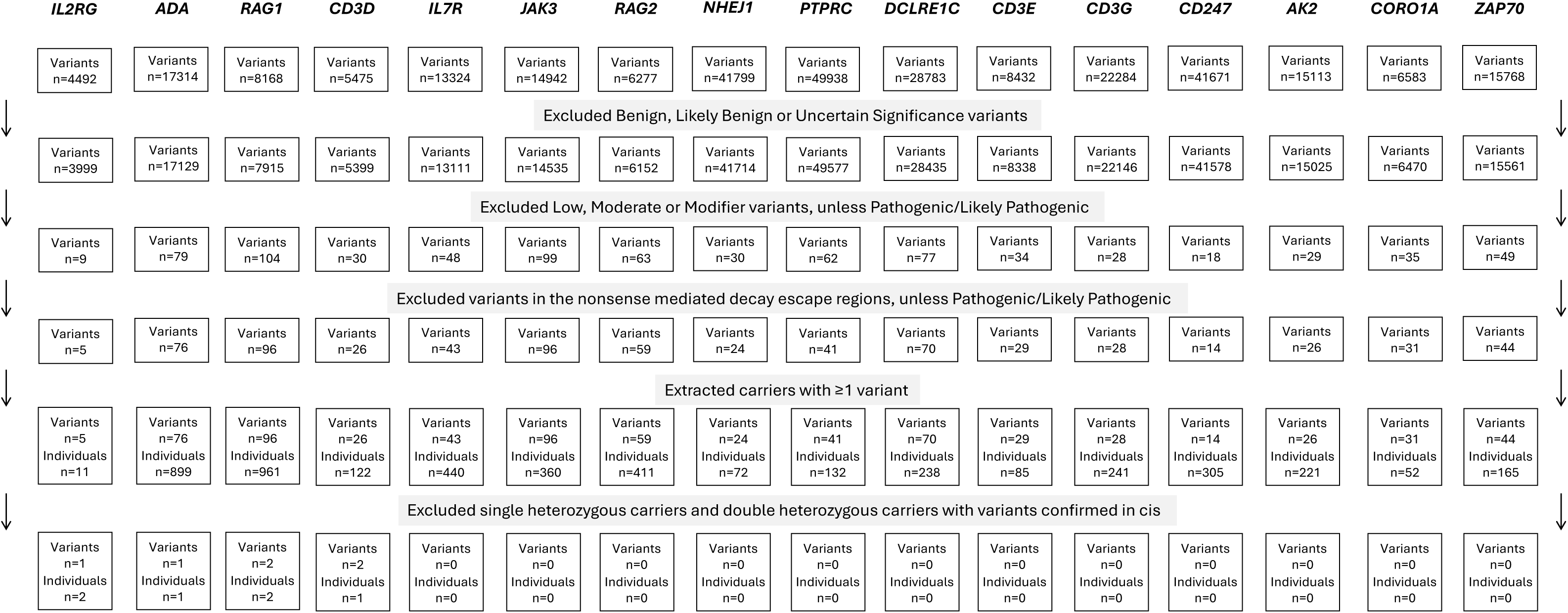
Flow diagram of the variant and genotype filtering strategy. Whole genome sequencing data from 490,640 individuals in the UK Biobank cohort, was interrogated for likely disease causing variation across 16 severe combined immunodeficiency genes. Variants with Benign, Likely Benign or Uncertain Significance ClinVar classifications were excluded (22). Variants with Variant Effect Predictor annotations of Low, Moderate or Modifier, were excluded unless they had Pathogenic/Likely Pathogenic ClinVar classifications. Variants in the nonsense mediated decay escape regions were excluded, unless Pathogenic/Likely Pathogenic or unless Pathogenic/Likely Pathogenic variants were present in these regions. Carriers of one or more variant were then extracted and homozygous, hemizygous and double heterozygous variant calls were visually assessed using the Integrative Genomics Viewer. Single heterozygous carriers and double heterozygous carriers with variants confirmed to be in cis were then excluded, leaving only individuals with potential biallelic pathogenic variation.

Variants were further excluded from analysis if there was no possibility of an individual in UKB carrying a biallelic genotype, i.e. being either homozygous (or hemizygous) for the variant or carrying ≥2 pathogenic heterozygous variants. Individual-level data for the remaining variants was manually inspected using the Integrative Genomics Viewer to verify variant calls. For heterozygous and hemizygous variants, those with a variant allele frequency less than 30% were excluded, given the likelihood of this indicating mosaic, rather than germline variation (24).

Phasing of double heterozygous variant carriers was carried out where variants were close enough to view simultaneously on the Integrative Genomics Viewer, using short-read sequencing data. Where possible, the gnomAD v2.1.1 co-occurrence data was used to conduct statistical phasing (25).

### Classifying the SCID phenotype

Classification of the SCID phenotype was approached using a strict and permissive eligibility criteria. The strict category contained only ICD-10 codes specific to SCID, which are D81.0, D81.1 and D81.2, and included two individuals in UKB. Due to the variability in SCID presentations and overlap with other immunodeficiencies (1), the permissive category was very broad (Supplementary Figure 1), including 656 participants. Subsequently, manual review was required to interpret phenotypes of homozygous, double heterozygous or hemizygous individuals.

## Results

### Lack of definitive biallelic pathogenic variants causing SCID in UKB

Across 16 genes, 4706 single heterozygous carriers were identified in UKB (∼1% of the cohort), with counts for each gene detailed in Figure 1. Potentially biallelic pathogenic variation was uncovered in 4/16 SCID genes, across 6 participants (Table 1), none of whom possessed an apparent SCID phenotype.

**Table 1.** Variants identified in individuals with potentially biallelic pathogenic variation.

| Gene (MANE select) | Variant ID (GRCh38) | HGVS | Location | Consequence | ClinVar Classification | Number of carriers in UKB | SCID diagnosis |
| --- | --- | --- | --- | --- | --- | --- | --- |
| *IL2RG*<br>(ENST00000374202.7) | X:71110236:G:T | c.514C>A<br>(p.Leu172Met) | Exon 4/8 | Missense variant | Likely Pathogenic (1*) | Male hemizygous: 2<br>Heterozygous: 4 | NA |
| *ADA*<br>(ENST00000372874.9) | 20:44625580:C:T | c.467G>A<br>(p.Arg156His) | Exon 5/12 | Missense variant | Pathogenic/Likely Pathogenic (2*) | Homozygous: 1 (VAF 88-89%)<br>Heterozygous: 33 | NA |
| *RAG1*<br>(ENST00000299440.6) | 11:36574870:G:T | c.1566G>T<br>(p.Trp522Cys) | Exon 2/2 | Missense variant | Pathogenic (2*) | Double heterozygous with c.2918G>A: 2<br>Total heterozygous: 124 | NA |
| *RAG1*<br>(ENST00000299440.6) | 11:36576222:G:A | c.2918G>A<br>(p.Arg973His) | Exon 2/2 | Missense variant | Pathogenic/Likely Pathogenic (2*) | Double heterozygous with c.1566G>T: 2<br>Total heterozygous: 3 | NA |
| *CD3D*<br>(ENST00000300692.9) | 11:118339871:CG:C | c.309del<br>(p.Val104TrpfsTer40) | Exon 3/5 | Frameshift variant | NA | Double heterozygous with c.202C>T: 1<br>Total heterozygous: 1 | NA |
| *CD3D*<br>(ENST00000300692.9) | 11:118340447:G:A | c.202C>T<br>(p.Arg68Ter) | Exon 2/5 | Stop gained | Pathogenic (2*) | Double heterozygous with c.309del: 1<br>Total heterozygous: 79 | NA |

Two male individuals were hemizygous for the ENST00000374202.7(*IL2RG*):c.514C>A (p.Leu172Met) variant, which had been retained due to a 1-star ClinVar Likely Pathogenic classification (22). No ICD-9, ICD-10 or GP codes were indicative of immunodeficiency, although one participant had IL2RG protein expression below the lower quartile for the cohort. However, on manual review following the ACMG/AMP guidelines (26), it was concluded that this was a variant of uncertain significance, primarily due its lack of identification in any affected individuals (22). This suggests that rather than being a low penetrance pathogenic variant, it is likely not disease-causing, providing an explanation for the lack of apparent phenotype in these two individuals.

One participant was called as homozygous for the ENST00000372874.9 (*ADA*):c.467G>A (p.Arg156His) variant, which has a 2-star Pathogenic/Likely Pathogenic ClinVar classification (22). However, manual review of sequencing reads revealed a REF/ALT ratio of 3/24 (variant allele fraction, VAF = 89%), with a similar VAF of 88% in exome sequencing data, likely caused by a somatic copy number loss over this locus due to blood cancer.

Two unrelated participants were identified with the same two missense variants in *RAG1*: ENST00000299440.6(*RAG1*):c.1566G>T (p.Trp522Cys) and c.2918G>A (p.Arg973His), which both have 2-star Pathogenic/Likely Pathogenic classifications on ClinVar (22). These two variants have not yet been reported in *trans* in a SCID patient. Owing to the distance between the variants, it was not possible to phase them directly using short-read WGS. However, the variants are predicted 100% likely to be in *trans* based on statistical phasing using the gnomAD co-occurrence data (25). The absence of SCID phenotypes in both individuals may therefore indicate that these are incompletely penetrant variants.

Finally, one participant was identified as double heterozygous for the variants ENST00000300692.9(*CD3D*):c.202C>T (p.Arg68Ter) and c.309del (p.Val104TrpfsTer40). The first variant was retained due to a 2-star Pathogenic ClinVar classification (22), while the second, which has not previously been recorded in a SCID patient, was retained due to a pLOF annotation. As with the *RAG1* variants, the variants could not be directly phased using short-read WGS; however, unlike the *RAG1* variants, statistical phasing was also not possible due to the absence of the novel variant in gnomAD. Consequently, it is not possible to discern whether these variants are in *trans* and exhibiting reduced penetrance, or were inherited in *cis* on the same allele.

### Low prevalence of SCID in adult population cohorts

While the incidence of SCID is estimated as 1 in 22,819 - 127,000, the prevalence in adult populations worldwide is unknown (8–10,27). Two individuals in UKB had specific SCID ICD-10 codes, indicating a prevalence of ∼1 in 250,000. One individual had several SCID ICD-10 codes, as well as various SCID-related healthcare records, although they also had ICD-10 codes indicative of other immunodeficiency disorders. The other participant had only one SCID ICD-10 code and no other reports that would be suggestive of SCID. As such there is a possibility that one or both of these diagnoses could have been miscoded; if so, prevalence may be as low, or even lower than ∼1 in 500,000 individuals.

None of the individuals potentially harbouring biallelic pathogenic variation had any SCID diagnoses recorded, nor any alternative immunodeficiency diagnoses. Additionally, the two reported SCID cases in UKB, were not found to have any potentially causative variation in the 16 genes analysed. As such, while the prevalence of causative variation in SCID genes is likely incredibly low, it was not possible to directly estimate it in this cohort.

## Discussion

The case for newborn screening for SCID is well supported, given the considerable improvements to survival that can be made with early initiation of HSCT (2–4). However, with the advent of gNBS, evidence is lacking as to how pathogenic SCID variants will be interpreted in an asymptomatic setting, with particular concerns surrounding reduced penetrance and variable expressivity. In the UKB, around 1% of the cohort carry a single heterozygous pathogenic variant in one of 16 SCID genes, whilst only 6 individuals (∼0.001% of the cohort) had potentially biallelic pathogenic variants, highlighting the importance of limiting variant reporting to double heterozygous and homozygous/hemizygous carriers. Upon further detailed curation, 3/6 of these individuals could be excluded based on poor analytical or clinical variant validity; the remaining 3/6 were double heterozygous carriers, of whom two were likely to be compound heterozygotes, but none of whom exhibited any SCID-related phenotypes.

Given the lack of any SCID phenotypes in the two individuals with likely compound heterozygous pathogenic variants, which is suggestive of incomplete penetrance, such findings should be interpreted with caution in asymptomatic newborns. While the p.Trp522Cys *RAG1* variant has not been previously reported in *trans* with p.Arg973His, as appears likely in these UKB individuals, it has been identified as compound heterozygous in an affected one-year-old with a variant in the same residue (p.Arg973Cys) (28). The fact that this highly similar genotype has previously been disease causing, provides further evidence to suggest that the genotype seen in the current study may be incompletely penetrant, potentially due to the hypomorphic nature of the p.Trp522Cys variant, which has been associated with 50% residual RAG1 protein activity (29). Although the p.Arg973His variant has been associated with less than 1% RAG1 activity (30), it is possible that the residual activity afforded by the p.Trp522Cys variant has prevented the development of SCID in these two individuals, hence the potential incomplete penetrance. Additionally, the p.Trp522Cys variant has been previously shown to exhibit variable expressivity (29,31–33). It has been identified in patients with severe, early onset SCID phenotypes, as compound heterozygous with other missense or nonsense variants (31,32). However, some p.Trp522Cys compound heterozygotes, have displayed milder phenotypes, with one individual presenting later in childhood despite the presence of a frameshift variant on the second allele, and another with an alternative missense variant surviving into adulthood without HSCT (29,33). As such, where hypomorphic variants are identified in an asymptomatic newborn, it is essential that they are interpreted with caution as they may exhibit incomplete penetrance or variable expressivity, even where pLOF or minimal activity variants are identified on the second allele. Given the recurrent nature of the p.Trp522Cys variant and its association with severe phenotypes (31,32), reporting in a gNBS setting may still be appropriate, although careful counselling will be required due to the uncertainty surrounding the clinical course, particularly if identified in a novel genotype. Given this uncertainty, while follow-up testing and monitoring of symptom onset may be advisable, presymptomatic treatment should be approached with utmost caution.

To limit the potential risk of overdiagnosis as a result of incomplete penetrance or erroneous variant interpretation, an essential prerequisite for gNBS programmes is suitable follow-up testing. Following a suspected-SCID gNBS result, this may involve genetic testing of parents to confirm phasing, measurement of enzyme activity, and lymphocyte phenotyping with flow cytometry (1). However, minimising the proportion of individuals requiring this secondary testing is essential, to limit undue stress for families, as well as excessive demand on paediatric immunologists and laboratories. In this cohort of 490,640, it is likely that only ∼3 individuals would require follow-up testing, had these variants been discovered in asymptomatic newborns, translating to fewer than 1 per 100,000. This finding corroborates several previous gNBS studies, which have identified no false positives resulting from variation in SCID genes (19,34–36). However, initial findings from the Generation Study, which has so far screened ∼30,000 newborns, have indicated that this false positive rate might be slightly higher, with two false positives having arisen due to variants in *ADA* and *RAG1*, respectively (37). Nonetheless, this indicates that the specificity of gNBS for SCID, likely exceeds that of TREC screening, for which a reported 16 in 100,000 individuals require follow-up testing (6). As TRECs are a specific marker for T-cell lymphopenia, rather than SCID, these SCID false positives may lead to alternative diagnoses, some of which have limited clinical actionability (10). This apparent specificity of gNBS for SCID therefore appears to confer a benefit over TREC screening, both in terms of minimising the need for follow up testing and in reducing alternative diagnoses.

However it is likely that this will largely depend on which genes are included for analysis by various gNBS programmes, particularly as certain SCID gene associations are less well established (1). Certain genes included in the Generation Study are poorly represented in other gNBS programmes, such as *CORO1A*, *CD3G, CD247* and *PTPRC*, likely due to the fact that there are very few case studies validating their causal role in SCID (13,18,19,35,36). Consequently it remains unclear whether variants in uncertain SCID genes may lead to false positives in newborns, although the lack of causative variation in the aforementioned genes in the UKB cohort, suggests that this should be of limited concern.

In addition to the importance of specificity, it is also essential that gNBS for SCID has appropriate sensitivity, to limit the number of missed cases. In our analysis of the 16 genes included in the Generation Study, we were unable to identify the causative variant(s) in the two individuals with SCID in the UKB, suggesting that WGS may lack complete sensitivity to detect SCID in newborns. It is possible that the lack of genetic findings in these individuals is due to the limited interrogation of copy number variation, or due to the difficulty interpreting novel missense or non-coding variation. In a newborn screening setting, variants without pLOF annotations or previous pathogenic assertions are unlikely to be further analysed, meaning that novel pathogenic missense or non-coding variants are likely to be overlooked (15). Given these individuals’ age and the unavailability of HSCT at the time of their births, it is possible that they possess a milder phenotype caused by novel hypomorphic variation, which was missed by this variant interpretation strategy (38). Another explanation for the lack of identified causal variation, is that these individuals possess causal variants in alternative SCID genes, beyond the 16 genes analysed in this study (1). However, while incorporation of an increasing number of genes in gNBS panels may improve sensitivity, the additional time required for variant interpretation may be unfeasible, particularly given that approximately 78% of SCID cases arise from variants in just 6 genes (1). As such, the lack of apparent causal variation in these two UKB individuals, raises concerns that some SCID cases may be missed by gNBS, whether due to novel variants or variants outside of specific gene panels. However, given that severe SCID phenotypes are typically caused by null variants which are easier to interpret, and that a large proportion of SCID arises from variants in only a few genes, it is expected that the majority of these severe cases will be identified by gNBS (1). Larger gNBS studies will likely provide more insight into whether cases will be missed due to the aforementioned limitations, and into how genomic analysis and variant interpretation can be improved to mitigate this in future gNBS programmes.

Additionally, one or both of these SCID cases may be the result of inaccurate clinical coding. One individual possessed a single SCID ICD-10 code and no other relevant clinical features, so it is plausible that their inclusion in this study is a result of miscoding. The other individual had multiple SCID ICD-10 codes alongside multiple ICD-10 codes for other immunodeficiencies, including “Other combined immunodeficiencies” and “Common variable immunodeficiency with autoantibodies to B- or T-cells”. These clinical codes are distinct from SCID and are often indicative of phenotypes presenting in adulthood (39). Furthermore, one study investigating hospital admissions for primary immunodeficiencies, found that due to a lack of distinction between “combined immune deficiency” and SCID in the historic ICD-9 coding system, only 15% of individuals with this code were truly likely to have SCID (40). It is possible that this historic lack of distinction has had a carry-over effect on the application of ICD-10 codes, or has led to difficulty in mapping ICD-9 to ICD-10. This is particularly plausible given that this individual possesses codes reflecting both of these previously conflated phenotypes. This altogether indicates that this participant may possess an alternative immunodeficiency phenotype which may be associated with variation in other genes. Consequently, as these individuals do not have conclusive SCID diagnoses, the lack of identified causal variation may be less indicative of limited sensitivity, potentially alleviating concerns about the sensitivity of WGS to detect SCID in gNBS.

This study had several limitations. Most importantly, the very low prevalence of biallelic pathogenic variant carriers seen in UKB may reflect the known healthy volunteer bias of this cohort, as well as the age of the participants (41). With the advent of potentially curative HSCT in the 1970-1980s, that was not available at the time of UKB participants’ births, it is likely that prevalence in younger populations is higher than the estimate provided here (2,38,42,43).

Consequently, large cohorts with younger individuals may be less depleted for SCID-causing variation, and will therefore provide more insight on population prevalence and penetrance. Long-read or trio sequencing will also be necessary to allow for definitive phasing and characterisation of compound heterozygous individuals, which was not possible in the current study. Furthermore, the accuracy of prevalence estimates is limited by the accuracy of clinical coding in the UKB, as well the lack of childhood healthcare records, that precluded confirmation of the early-onset phenotype that is characteristic of SCID. It is also possible that potential compound heterozygotes in fact possessed a SCID diagnosis, that had not been accurately recorded, although none of these participants possessed any broader immunodeficiency phenotypes. With the increase in genetic diagnoses for SCID patients afforded by gNBS and the standardisation of genetic testing for suspected cases, it is likely that the accuracy of SCID clinical coding will improve, aiding future investigations of penetrance and prevalence.

## Conclusion

The absence of confirmed biallelic pathogenic variation in 16 SCID genes, in almost 500,000 unaffected individuals, is strongly suggestive of high penetrance. This indicates that false positives should be of limited concern in gNBS for SCID, particularly given the minimal number of individuals in the UKB that would require follow-up testing if identified in a gNBS setting.

However, the relatively high single heterozygous carrier rate of ∼1% highlights the importance of focusing on double heterozygous or homozygous/hemizygous carriers. In addition, the lack of apparent phenotypes in the two individuals likely to be compound heterozygous carriers, suggests that caution is needed when reporting novel combinations of potentially hypomorphic missense variants in asymptomatic newborns. Nonetheless, while the consequences of reporting hypomorphic missense variants must be considered, SCID remains an exemplary candidate for gNBS, given its severity, clinical actionability and ostensibly high penetrance.

## Data and code availability statement

Data from UK Biobank is not publicly available because of data governance and data return policies. Data are available for researchers who meet the criteria and receive approval to access the UK Biobank dataset (www.ukbiobank.ac.uk).

## Data Availability

Data from UK Biobank is not publicly available because of data governance and data return
policies. Data are available for researchers who meet the criteria and receive approval to
access the UK Biobank dataset (www.ukbiobank.ac.uk).

## Acknowledgments and funding statement

We gratefully acknowledge participants of the UK Biobank for their contributions, without which this research would not have been possible. This research has been carried out using the UK Biobank Resource under Application Number 103356, and includes data provided by patients and collected by the National Health Service as part of their care. The authors would like to acknowledge the use of the University of Exeter High-Performance Computing (HPC) facility in facilitating this work.

This work was supported by the Medical Research Council [MR/X021351/1] and the National Institute for Health and Care Research Exeter Biomedical Research Centre. The views expressed are those of the author(s) and not necessarily those of the NIHR or the Department of Health and Social Care.

## Author Contribution Statement

IJHG- Investigation, Visualisation, Writing- original draft preparation, Writing – review & editing JF- Investigation, Writing – review & editing

CFW- Funding curation, Conceptualisation, Writing – review & editing

LJ- Funding curation, Conceptualisation, Data Curation, Supervision, Writing – review & editing

## Ethical Approval

UK Biobank protocols were approved by the National Research Ethics Service Committee.

## References

1. Dvorak CC, Haddad E, Heimall J, Dunn E, Buckley RH, Kohn DB, et al. The diagnosis of severe combined immunodeficiency (SCID): The Primary Immune Deficiency Treatment Consortium (PIDTC) 2022 Definitions. J Allergy Clin Immunol. 2023 Feb;151(2):539–46.

2. Lankester AC, Neven B, Mahlaoui N, von Asmuth EGJ, Courteille V, Alligon M, et al. Hematopoietic cell transplantation in severe combined immunodeficiency: The SCETIDE 2006-2014 European cohort. J Allergy Clin Immunol. 2022 May;149(5):1744–1754.e8.

3. Haddad E, Logan BR, Griffith LM, Buckley RH, Parrott RE, Prockop SE, et al. SCID genotype and 6-month posttransplant CD4 count predict survival and immune recovery. Blood. 2018 Oct 25;132(17):1737–49.

4. Heimall J, Logan BR, Cowan MJ, Notarangelo LD, Griffith LM, Puck JM, et al. Immune reconstitution and survival of 100 SCID patients post–hematopoietic cell transplant: a PIDTC natural history study. Blood. 2017 Dec 21;130(25):2718–27.

5. Brown L, Xu-Bayford J, Allwood Z, Slatter M, Cant A, Davies EG, et al. Neonatal diagnosis of severe combined immunodeficiency leads to significantly improved survival outcome: the case for newborn screening. Blood. 2011 Mar 17;117(11):3243–6.

6. Amatuni GS, Currier RJ, Church JA, Bishop T, Grimbacher E, Nguyen AAC, et al. Newborn Screening for Severe Combined Immunodeficiency and T-cell Lymphopenia in California, 2010–2017. Pediatrics. 2019 Feb;143(2):e20182300.

7. Currier R, Puck JM. SCID newborn screening: What we’ve learned. Journal of Allergy and Clinical Immunology. 2021 Feb;147(2):417–26.

8. Liao HC, Liao CH, Kao SM, Chiang CC, Chen YJ. Detecting 22q11.2 Deletion Syndrome in Newborns with Low T Cell Receptor Excision Circles from Severe Combined Immunodeficiency Screening. J Pediatr. 2019 Jan;204:219–24.e1.

9. Booth NA, Freeman CM, Wright BL, Rukasin C, Badia P, Daines M, et al. Severe Combined Immunodeficiency (SCID) Screening in Arizona: Lessons Learned from the First 2 Years. J Clin Immunol. 2022 Aug 22;42(6):1321–9.

10. Kleijnen Systematic Reviews Ltd. Newborn screening for severe combined immunodeficiency (SCID) in the NHS Newborn Blood Spot (N B S) screening programme: A rapid evidence review. UK National Screening Committee. 2025. https://nationalscreening.blog.gov.uk/wp-content/uploads/sites/254/2025/07/UK-NSC-Newborn-screening-for-SCID-evidence-summary-July-2025.pdf. Accessed 10 Dec 2025.

11. Genomics England. Genomics England PanelApp [Internet]. [cited 2025 Dec 15]. https://panelapp.genomicsengland.co.uk/

12. Kohn DB, Hershfield MS, Puck JM, Aiuti A, Blincoe A, Gaspar HB, et al. Consensus approach for the management of severe combined immune deficiency caused by adenosine deaminase deficiency. J Allergy and Clin Immunol. 2019 Mar;143(3):852–63.

13. Minten T, Bick S, Adelson S, Gehlenborg N, Amendola LM, Boemer F, et al. Data-driven consideration of genetic disorders for global genomic newborn screening programs. Genet Med. 2025 Jul;27(7):101443.

14. Freeman K, Taylor D, Dinnes J, Clark CCA, Kander I, Scandrett K, et al. Challenges in evaluating whole genome sequencing for newborn screening: series of systematic reviews and roadmap for evidence generation for policy advisers. BMJ Med. 2025 Nov 21;4(1):e001726.

15. Kaplanis J, Deen D, Sivakumar P, de Almeida Gomes M, Puzriakova A, Leong I, et al. Assessment of the variant prioritization strategy for genomic newborn screening in the Generation Study. Genet Med. 2025 Oct;27(10):101532.

16. Forrest IS, Chaudhary K, Vy HMT, Petrazzini BO, Bafna S, Jordan DM, et al. Population-Based Penetrance of Deleterious Clinical Variants. JAMA. 2022 Jan 25;327(4):350–9.

17. Wright CF, Sharp LN, Jackson L, Murray A, Ware JS, MacArthur DG, et al. Guidance for estimating penetrance of monogenic disease-causing variants in population cohorts. Nat Genet. 2024 Sep;56(9):1772–9.

18. Ceyhan-Birsoy O, Machini K, Lebo MS, Yu TW, Agrawal PB, Parad RB, et al. A curated gene list for reporting results of newborn genomic sequencing. Genet Med. 2017 Jul;19(7):809–18.

19. Ziegler A, Koval-Burt C, Kay DM, Suchy SF, Begtrup A, Langley KG, et al. Expanded Newborn Screening Using Genome Sequencing for Early Actionable Conditions. JAMA. 2025 Jan 21;333(3):232–40.

20. UK Biobank. [cited 2025 Dec 20]. https://www.ukbiobank.ac.uk

21. Carss K, Halldorsson B V., Hou L, Liu J, Wheeler E, Lo Y, et al. Whole-genome sequencing of 490,640 UK Biobank participants. Nature. 2025 Sep;645(8081):692–701.

22. ClinVar. [cited 2025 Mar 22]. https://www.ncbi.nlm.nih.gov/clinvar/

23. Khajavi M, Inoue K, Lupski JR. Nonsense-mediated mRNA decay modulates clinical outcome of genetic disease. Eur J Hum Genet. 2006 Oct ;14(10):1074–81.

24. Coffee B, Cox HC, Kidd J, Sizemore S, Brown K, Manley S, et al. Detection of somatic variants in peripheral blood lymphocytes using a next generation sequencing multigene pan cancer panel. Cancer Genet. 2017 Feb;211:5–8.

25. Karczewski KJ, Francioli LC, Tiao G, Cummings BB, Alföldi J, Wang Q, et al. The mutational constraint spectrum quantified from variation in 141,456 humans. Nature. 2020 May ;581(7809):434–43.

26. Richards S, Aziz N, Bale S, Bick D, Das S, Gastier-Foster J, et al. Standards and guidelines for the interpretation of sequence variants: a joint consensus recommendation of the American College of Medical Genetics and Genomics and the Association for Molecular Pathology. Genet Med. 2015 May;17(5):405–24.

27. Orphanet. [cited 2025 Dec 23]. https://www.orpha.net/en/disease/detail/183660

28. ID bases. Rag1base. [cited 2026 Jan 27]. https://structure.bmc.lu.se/idbase/RAG1base/index.php?content=pub/IDbases

29. De Ravin SS, Cowen EW, Zarember KA, Whiting-Theobald NL, Kuhns DB, Sandler NG, et al. Hypomorphic Rag mutations can cause destructive midline granulomatous disease. Blood. 2010 Aug 26;116(8):1263–71.

30. Corneo B, Moshous D, Güngör T, Wulffraat N, Philippet P, Deist F Le, et al. Identical mutations in RAG1 or RAG2 genes leading to defective V(D)J recombinase activity can cause either T-B–severe combined immune deficiency or Omenn syndrome. Blood. 2001 May 1;97(9):2772–6.

31. Villa A, Sobacchi C, Notarangelo LD, Bozzi F, Abinun M, Abrahamsen TG, et al. V(D)J recombination defects in lymphocytes due to RAG mutations: severe immunodeficiency with a spectrum of clinical presentations. Blood. 2001 Jan 1;97(1):81–8.

32. Meng L, Pammi M, Saronwala A, Magoulas P, Ghazi AR, Vetrini F, et al. Use of Exome Sequencing for Infants in Intensive Care Units. JAMA Pediatr. 2017 Dec 4;171(12):e173438.

33. Csomos K, Ujhazi B, Blazso P, Herrera JL, Tipton CM, Kawai T, et al. Partial RAG deficiency in humans induces dysregulated peripheral lymphocyte development and humoral tolerance defect with accumulation of T-bet+ B cells. Nat Immunol. 2022 Aug;23(8):1256–72.

34. Ceyhan-Birsoy O, Murry JB, Machini K, Lebo MS, Yu TW, Fayer S, et al. Interpretation of Genomic Sequencing Results in Healthy and Ill Newborns: Results from the BabySeq Project. Am J Hum Genet. 2019 Jan 3;104(1):76–93.

35. Pavey AR, Bodian DL, Vilboux T, Khromykh A, Hauser NS, Huddleston K, et al. Utilization of genomic sequencing for population screening of immunodeficiencies in the newborn. Genet Med. 2017 Dec;19(12):1367–75.

36. Bodian DL, Klein E, Iyer RK, Wong WSW, Kothiyal P, Stauffer D, et al. Utility of whole-genome sequencing for detection of newborn screening disorders in a population cohort of 1,696 neonates. Genet Med. 2016 Mar;18(3):221–30.

37 . Bick D. Newborn Genome Screening: Initial results of the Generation Study. Presented at: The Festival of Genomics and Biodata; 28th Jan 2026; London, England.

38. Hobbs JR. Bone marrow transplantation for inborn errors. Lancet. 1981 Oct 3;2(8249):735–9.

39. Thaventhiran JED, Lango Allen H, Burren OS, Rae W, Greene D, Staples E, et al. Whole-genome sequencing of a sporadic primary immunodeficiency cohort. Nature. 2020 Jul;583(7814):90–5.

40. Resnick ES, Bhatt P, Sidi P, Cunningham-Rundles C. Examining the Use of ICD-9 Diagnosis Codes for Primary Immune Deficiency Diseases in New York State. J Clin Immunol. 2012 Sep 1;33(1):40–8.

41. Fry A, Littlejohns TJ, Sudlow C, Doherty N, Adamska L, Sprosen T, et al. Comparison of Sociodemographic and Health-Related Characteristics of UK Biobank Participants With Those of the General Population. Am J Epidemiol. 2017 Nov 1;186(9):1026–34.

42. Blom M, Bredius RGM, van der Burg M. Efficient screening strategies for severe combined immunodeficiencies in newborns. Expert Rev Mol Diagn. 2023 Jul;23(9):815–25.

43. Dorsey MJ, Wright NAM, Chaimowitz NS, Dávila Saldaña BJ, Miller H, Keller MD, et al. Infections in Infants with SCID: Isolation, Infection Screening, and Prophylaxis in PIDTC Centers. J Clin Immunol. 2021 Jan;41(1):38–50.

