## Supplementary Table 1 for "Investigating penetrance of severe combined immunodeficiency variants in an adult population cohort: implications for genomic newborn screening"

| ICD-9/10 | Code | Description |
|---|---|---|
| ICD-10 | D80 | Immunodeficiency with predominantly antibody defects |
| ICD-10 | D80.5 | Immunodeficiency with increased immunoglobulin in M (IgM) |
| ICD-10 | D80.6 | Immunodeficiency with predominantly antibody defects, with hyperimmunoglobulinemia or normal immunoglobulins |
| ICD-10 | D80.8 | Other immunodeficiency with predominantly antibody defects, specified type not elsewhere classified |
| ICD-10 | D80.9 | Immunodeficiency with predominantly antibody defect |
| ICD-10 | D81 | Combined immunodeficiencies |
| ICD-10 | D81.3 | Adenosine deaminase [ADA] deficiency |
| ICD-10 | D81.8 | Combined immunodeficiency, specified type not elsewhere classified |
| ICD-10 | D81.9 | Unspecified combined immunodeficiency, combined severe (SCID) |
| ICD-10 | D82 | Immunodeficiency with other major defects |
| ICD-10 | D82.2 | Immunodeficiency with short-limbed stature |
| ICD-10 | D82.8 | Immunodeficiency associated with major defect, specified type not elsewhere classified |
| ICD-10 | D82.9 | Immunodeficiency associated with major defect |
| ICD-10 | D83 | Common variable immunodeficiency |
| ICD-10 | D83.0 | Common variable immunodeficiency with abnormalities of B-cell numbers and function |
| ICD-10 | D83.1 | Common variable immunodeficiency with immunoregulatory T-cell disorders |
| ICD-10 | D83.2 | Common variable immunodeficiency with autoantibodies to B- or T-cells |

| | | |
|---|---|---|
| ICD-10 | D83.8 | Common variable immunodeficiency with specified type not classified elsewhere |
| ICD-10 | D83.9 | Common variable immunodeficiency, unspecified |
| ICD-10 | D84 | Other immunodeficiencies |
| ICD-10 | D84.8 | Immunodeficiency, specified type not elsewhere classified |
| ICD-10 | D84.9 | Immunodeficiency, unspecified |
| ICD-9 | 2772 | Other disorders of purine and pyrimidine metabolism |
| ICD-9 | 2790 | Deficiency of humoral immunity |
| ICD-9 | 27905 | Deficiency of humoral immunity (dysimmunoglobulinaemia) |
| ICD-9 | 27906 | Common variable immunodeficiency |
| ICD-9 | 27909 | Other and unspecified deficiency of humoral immunity |
| ICD-9 | 2791 | Deficiency of cell-mediated immunity |
| ICD-9 | 27919 | Other deficiency of cell-mediated immunity |
| ICD-9 | 2792 | Combined immunity deficiency |
| ICD-9 | 2793 | Unspecified immunity deficiency |
| ICD-9 | 2798 | Other specified disorders involving the immune mechanism |
| ICD-9 | 2799 | Other specified disorders involving the immune mechanism |

**Supplementary Table 1. List of ICD-9 and ICD-10 codes defining the permissive phenotype eligibility criteria.**
